# The landscape of secondary malignancies in cancer survivors and the susceptibility to hereditary multiple cancer syndrome: a Korean nationwide population-based study

**DOI:** 10.1101/2022.06.08.22276130

**Authors:** Yoon Young Choi, Myeongjee Lee, Eun Hwa Kim, Jae Eun Lee, Inkyung Jung, Jae-Ho Cheong

## Abstract

**Background:** Cancer survivors and secondary malignancy risk have increased following early diagnosis and advanced cancer treatments; however, secondary malignancy risk in cancer survivors and possibility of genetic susceptibility among young individuals with multiple cancers have scarcely been evaluated. We evaluated the risk of secondary malignancies in cancer survivors by primary cancer type, and the combination of cancer types frequently observed in young patients with multiple cancers.

**Methods and Findings:** Patients diagnosed with cancer, from Korea Health Insurance Review and Assessment database, between January 2009 and December 2010, were followed-up until December 2019. Cancer survivors and those with secondary cancers were defined as having lived >5 years without developing other cancers; and being newly diagnosed with cancer at another site, respectively. To identify possible hereditary multiple cancer syndrome (HMCS), combinations of multiple cancers frequently observed in young individuals compared to older adults were evaluated. Of the 371,181 patients in 3,008,274 person-years of follow-up, 266,241 were cancer survivors; of these 7,348 had secondary cancers (2.76%; 2,759.9 persons/100,000 population), higher than primary cancer risk in general population. The common primary cancer types were those mainly observed as secondary cancer in cancer survivors. Contrarily, higher-risk cancers among those with secondary cancers compared to those in the general population varied by sex and primary cancer type, suggesting the need for tailored cancer screening for cancer survivors. Combinations of well-known hereditary cancer syndrome-related cancer types were mainly observed in younger compared to older patients with multiple cancers, which could be HMCS, including cancer combinations those may be caused by a novel cancer predisposition gene.

**Conclusions:** Cancer survivors harbor high and distinctive secondary cancer risk than the general population, necessitating tailored cancer screening in survivors. Genetic screening should be considered for young patients with multiple cancers to validate genetic predisposition.

## INTRODUCTION

Cancer, a global health burden, is the main cause of death globally.[1] Early diagnosis and advanced treatment strategy have improved the survival of cancer patients; therefore, managing the health of cancer survivors, including those with secondary cancers, has become an important medical issue.[2, 3] Because cancer survivors live in similar environments as where their first cancer developed and the genetic risk is sustained, the risk of secondary cancer could be associated with the first one. Consequently, the landscape of secondary cancers may differ compared to the primary cancer in the general population. Therefore, tailored cancer screening and prevention by the type of first cancer is necessary. Although epidemiologic evidence has been reported on some single cancer types,[4-7] there has been no study focused on the overall association between the primary and secondary cancers, targeting cancer survivors so far.

Family history of cancer is the classic surrogate for hereditary cancer syndrome caused by genetic susceptibility. However, as life expectancy has increased, the possibility of experiencing cancer during one’s lifetime has increased. Consequently, only family history could be a sensitive but not specific surrogate to guide hereditary cancer syndrome. Along with younger-age onset cancer, multiple cancers are regarded as being related to hereditary cancer syndrome.[8, 9] A recent study reported that younger patients with multiple Lynch syndrome-related cancers showed a strong association with genetic susceptibility.[10] Therefore, the combination of multiple cancers frequently observed in younger individuals could be a gateway to guide hereditary multiple cancer syndrome (HMCS) (i.e., multiple cancers caused by genetic susceptibility). However, this has not been assessed in epidemiologic studies.

In this study, we evaluated the high-risk secondary malignancies in cancer survivors (SMCS) by primary cancer type and identified the candidates for HMCS, using data from the South Korea Health Insurance Review and Assessment (HIRA) database.

## METHODS

### Data source

This nationwide, population-based, cohort study was conducted using data collected in the HIRA database between January 2008 and December 2019, which includes over 50 million Koreans. This study was approved by the research ethics committee of South Korea National Health Insurance Sharing Service (M20200902739) and the institutional review board of Severance Hospital (4-2020-0155). The need for informed consent was waived owing to the use of de-identified data.

### Patient and Public Involvement

Patients or the public were not involved in the design, or conduct, or reporting, or dissemination plans of our research

### Study population

We retrieved the data of patients diagnosed with cancer between January 2009 and December 2010. To identify newly diagnosed cancer cases during this period, those diagnosed in 2008 were excluded. We used the 10^th^ revision of the International Classification of Diseases (ICD) code and the Korean Individual Co-payment Beneficiaries Program (ICBP) data to define each cancer site. The ICBP was established for rare and intractable diseases, including cancers, in 2008, and patients in this program pay only 5-10% of their medical costs. Therefore, the most affected individuals are registered in ICBP. We followed those cancer patients until the end of December 2019. Cancer survivors were defined as cancer patients who have lived longer than 5 years without developing other cancers besides the first diagnosed cancers. SMCS was defined as a newly diagnosed distinct cancer type after 5-years of the primary cancer diagnosis (cancer at the same site as the initial one was not considered). We hypothesized that the combination of cancer types frequently observed in younger compared to older individuals is related to hereditary cancer. To validate this hypothesis, cancer patients with HMCS were defined based on a diagnosis with specific combinations of cancer types and more often diagnosed at a younger age for the first cancer. Therefore, we collected all information on diagnosed cancer types in cancer patients during the study period and included patients with two or more cancers. We then defined the combinations of multiple cancer types and categorized younger and older age groups depending on their age at the first cancer diagnosis.

### Statistical analysis

To investigate a potentially increased risk of secondary cancer in cancer survivors compared to the general Korean population, we calculated the age-standardized incidence ratio (SIR) by primary cancer type and sex, using the Korean National Cancer Statistics 2015,[11] the median time point of the present cohort as the reference values. The 95% confidence interval (CI) of the SIR was calculated using the Poisson distribution. For each primary cancer, the high-risk SMCS was defined when the SIR was statistically significantly greater than 1.

To identify the HMCS candidates, the relative risk (RR) of each combination of cancers in younger compared to older individuals was calculated. For patients with two distinct cancer types, younger and older individuals were defined as being younger and older than 10-years from the median age at diagnosis for each first cancer,[12-15] respectively, by sex (S1 Table 1). Considering the complexity of combinations and small sample sizes of patients with three or more distinct cancers, the younger and older individuals were defined as being < and ≥50 years based on the age at first cancer diagnosis.[16, 17] The ratio was compared to that of patients with multiple cancers (n=60,648) by sex, and the order of cancer diagnosis was not considered. Combinations of cancers with over 5 cases were statistically analyzed. A two-sided p-value <0.05 was considered statistically significant. SAS Enterprise Guide version 9.4 (SAS Institute Inc., Cary, NC, USA) was used for all statistical analyses.

**Table 1.** Number and proportion of patients by primary cancer types in the study cohort

| ICD | Primary cancer | No. of patients | Proportion (%) |
| --- | --- | --- | --- |
| C00-14 | Head & Neck | 5,534 | 1.49 |
| C15 | Esophagus | 3,702 | 1.0 |
| C16 | Stomach | 61,775 | 16.64 |
| C18-20 | Colo-rectum | 49,614 | 13.37 |
| C22 | Liver | 28,858 | 7.77 |
| C23-24 | GB-biliary | 7,332 | 1.98 |
| C25 | Pancreas | 7,392 | 1.99 |
| C32 | Larynx | 2,468 | 0.66 |
| C33-34 | Lung | 32,182 | 8.67 |
| C50 | Breast | 34,706 | 9.35 |
| C53 | Cervix | 11,144 | 3.0 |
| C54 | Uterus | 3,642 | 0.98 |
| C56 | Ovary | 4,680 | 1.26 |
| C61 | Prostate | 13,539 | 3.65 |
| C62 | Testis | 493 | 0.13 |
| C64 | Kidney | 7,065 | 1.9 |
| C67 | Bladder | 8,592 | 2.31 |
| C70-72 | Brain | 5,254 | 1.42 |
| C73 | Thyroid | 69,211 | 18.65 |
| C81 | Hodgkin lymphoma | 455 | 0.12 |
| C82-86,<br>C96 | Non-Hodgkin lymphoma | 7,001 | 1.89 |
| C90 | Multiple myeloma | 1,942 | 0.52 |
| C91-95 | Leukemia | 4,600 | 1.24 |
| Total |  | 371,181 | 100 |

ICD; International Classification of Diseases, GB; gallbladder

## RESULTS

### Population

A total of 371,181 patients were diagnosed with primary cancers between 2009 and 2010 and followed up in 3,008,274 person-years. Table 1 shows the detailed number of enrolled patients by primary cancer types, and the proportions by cancer type were similar to those in Korean cancer statistics.[18] After excluding 76,103 deaths and 28,837 cases with additional cancers within 5 years, there were 266,241 cancer survivors and 7,348 of these (2.76%, 2,759.9 person/100,000 population) were diagnosed with secondary cancer. This rate is substantially higher than the most recent Korean cancer statistics in 2019; 475.3 person/100,000 population of incidence rate for primary cancer in the overall Korean population.[18]

There were 60,648 cancer patients diagnosed with two or more distinct cancer types during follow-up; 50,561 patients had two distinct cancers and 10,087 patients had three or more distinct cancers (Figure 1).

**Figure 1.**
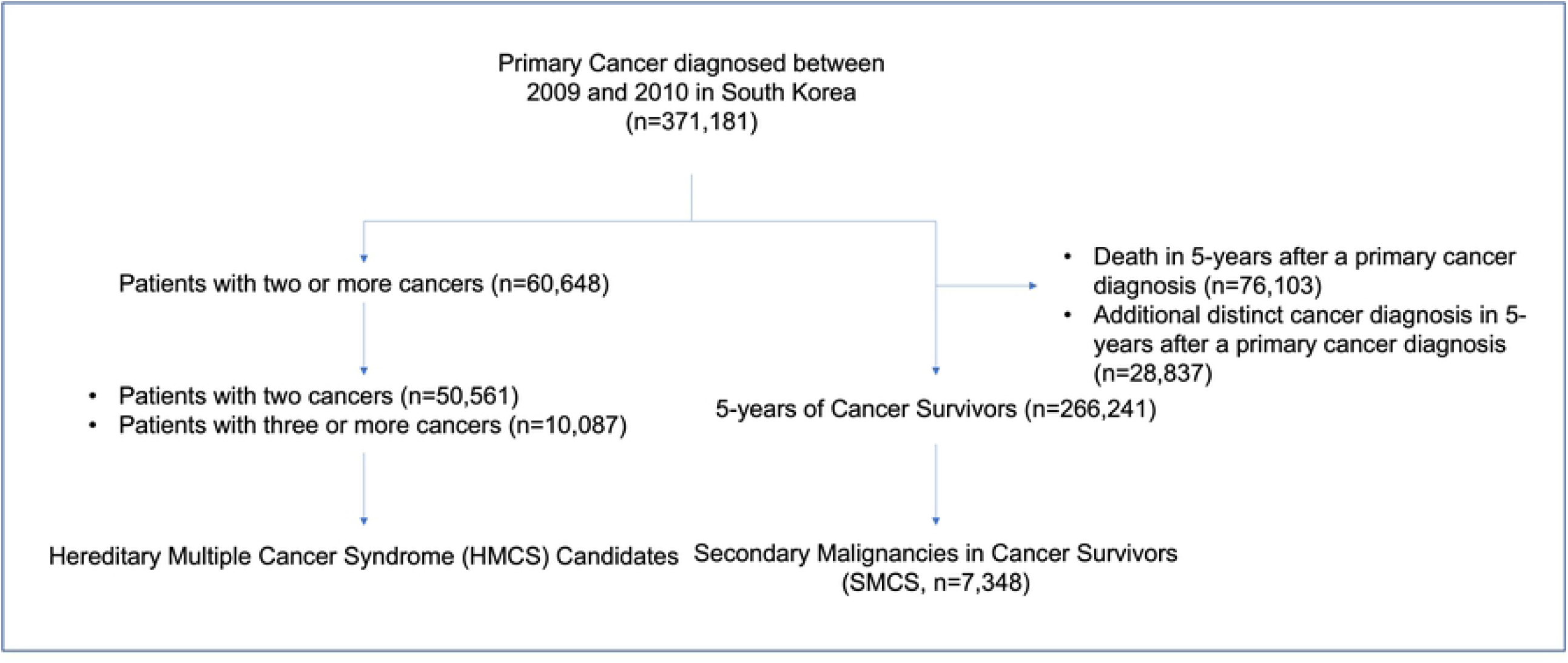
Flow diagram for patient selection.

### SMCS

The risk of secondary cancer was highest in laryngeal cancer survivors (9.03%, Table 2) and lowest in multiple myeloma survivors (0.63%). There was no significant correlation between the percentage of survivors according to cancer type and the risk of SMCS with cancer (pearson’s correlation p=0.512). In breast cancer survivors, the risk of secondary cancer was higher in male patients compared to female patients (p=0.002).

**Table 2.**
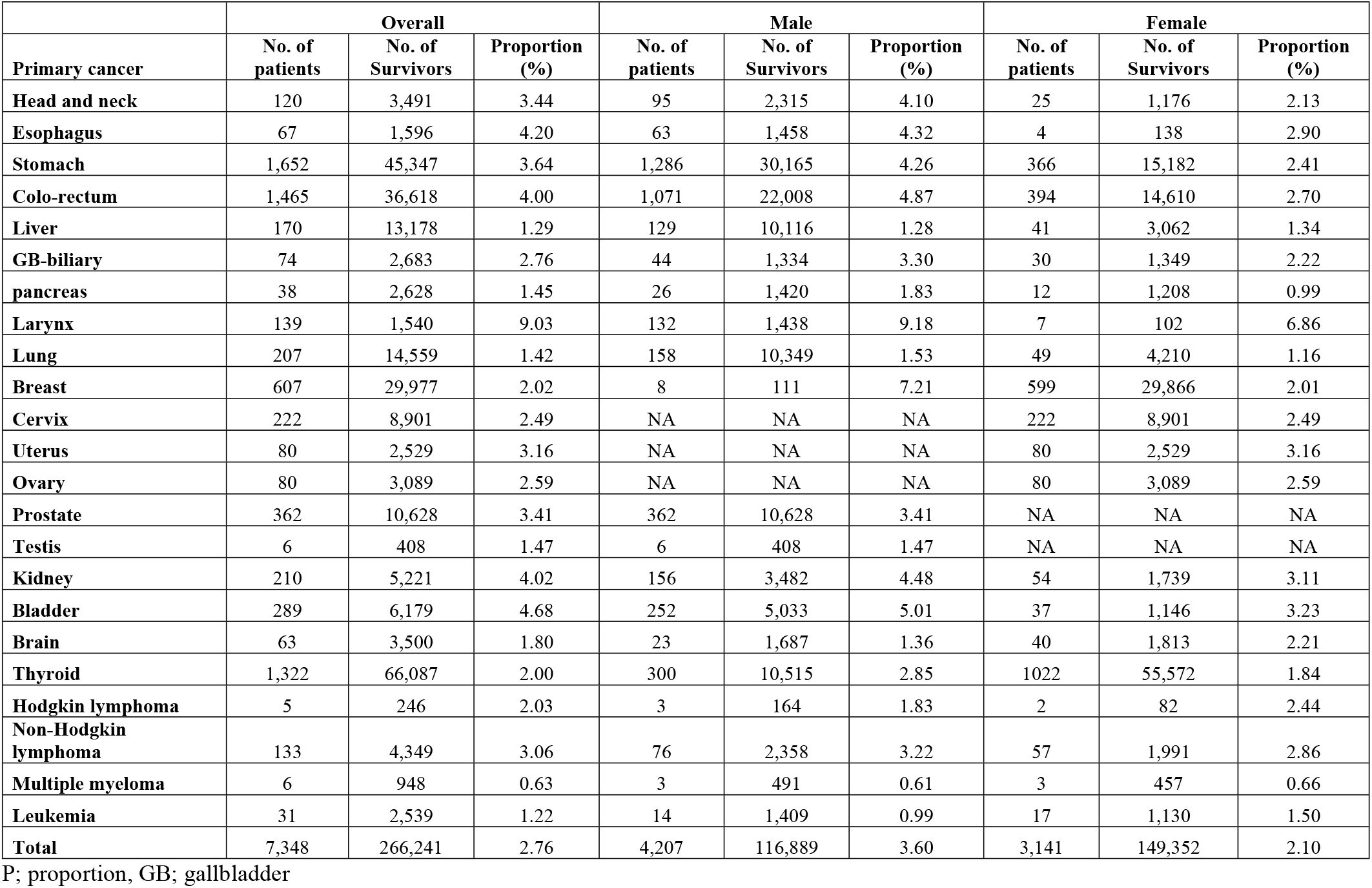
Number and proportion of patients with secondary cancers at other anatomical sites among cancer survivors

The most frequent SMCS were lung, prostate, stomach, colo-rectum, and liver cancers in male patients; and breast, lung, stomach, colo-rectum, and thyroid cancers in female patients (S1 Tables 2 and 3). These cancers were the most frequent primary cancers by sex in Korea.[18] Contrarily, the landscape of high-risk SMCS differed from that in the general population: the risk of prostate cancer was 7.931 times higher in male breast cancer survivors, while the risk of lung cancer was 10.093 times higher in female laryngeal cancer survivors compared to the general population (Figure 2A and B, S1 table 4 and 5). The frequently observed high-risk secondary cancers in male and female cancer survivors were prostate, bladder, and lung cancers; and pancreas and lung cancers, respectively. The risk of prostate cancer as a secondary cancer compared to that in the general population was increased in male survivors of stomach, colo-rectum, breast, kidney, and thyroid cancers, while the risk of pancreas cancer as a secondary cancer was increased in survivors of colo-rectum, breast, kidney, thyroid cancer, and non-Hodgkin lymphoma in female. In addition, some cancer types were highly associated with each other regardless of the order of diagnosis; in male patients, the risk of cancer of the head & neck/esophagus, and larynx/bladder were high (SIRs; 5.556/6.357 and 2.657/2.698, respectively, Figure 2A), suggesting that smoking could affect even secondary cancers because these are all smoking-related cancers.[19] The risk of cancer of the breast/uterus (possibly due to hormonal effect) and non-Hodgkin lymphoma/leukemia (possibly due to chemotherapy effect),[20] was high in female patients (SIRs; 2.82/1.757 and 7.05/8.553, respectively, Figure 2B).

**Table 3.**
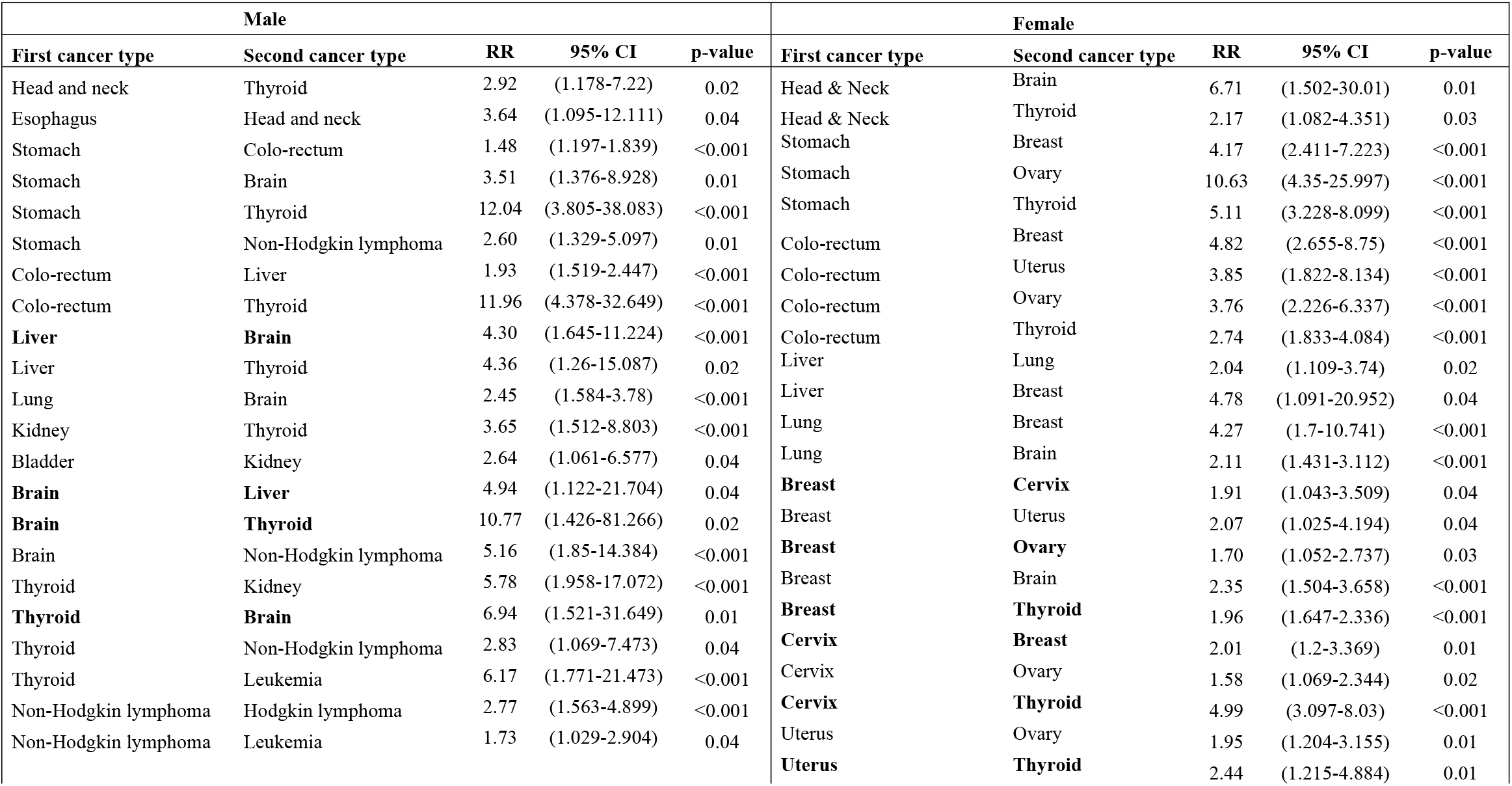
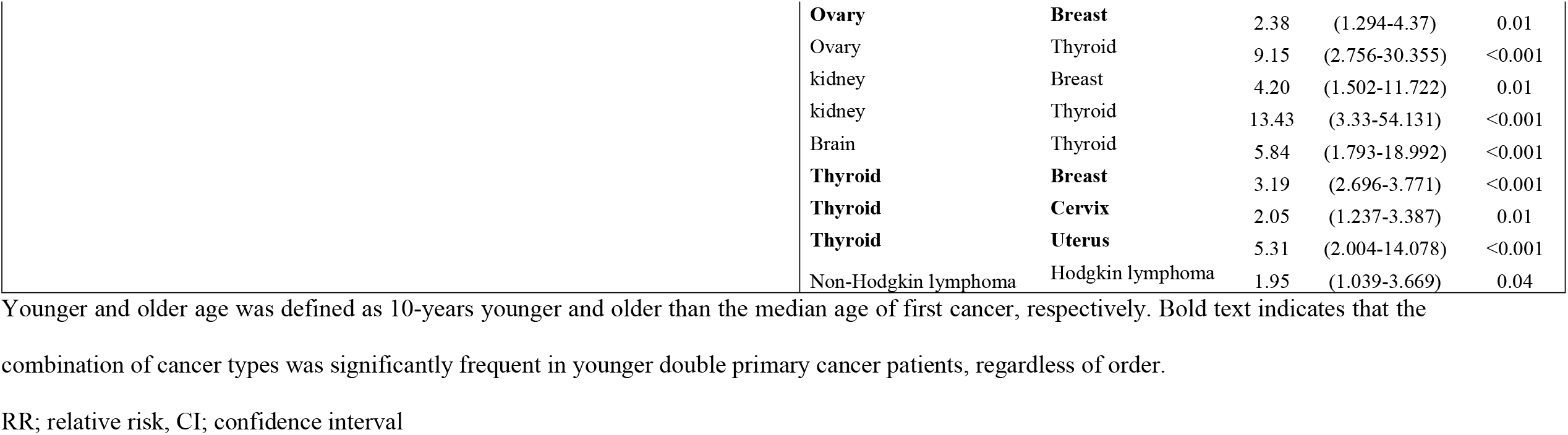
Relative risk of combination of double primary cancers in younger compared to older patients

**Table 4.** High-risk combination of multiple cancers in younger (<50 years old) compared to older (≥50 years) patients with three or more primary cancers

| | Cancer types<sup>†</sup> | No. of distinct cancer types | No. of patients < 50years (%)<sup>‡</sup> | No. of patients $\geq 50$ years (%) | RR (95% CI) * | p-value |
| --- | --- | --- | --- | --- | --- | --- |
| **Male** | Colo-rectum/Liver/GB-biliary/Pancreas | 4 | 8 (20.5) | 31 (79.5) | 2.473 (1.138-5.376) | 0.027 |
|  | Stomach/Colo-rectum/Pancreas | 3 | 36 (16.9) | 177 (83.1) | 1.949 (1.365-2.784) | 0.001 |
|  | Hodgkin/Non-Hodgkin lymphoma/Leukemia | 3 | 7 (63.6) | 4 (36.4) | 16.771 (4.912-57.261) | <0.001 |
|  | Pancreas/Prostate/Brain | 3 | 7 (41.2) | 10 (58.8) | 6.709 (2.555-17.612) | 0.001 |
|  | Liver/Lung/Brain | 3 | 6 (25.0) | 18 (75.0) | 3.195 (1.269-8.042) | 0.021 |
| **Female** | Colo-rectum/Cervix/Uterus/Ovary | 4 | 7 (70.0) | 3 (30.0) | 6.162 (1.594-23.822) | 0.006 |
|  | Colo-rectum/Pancreas/Uterus/Ovary | 4 | 6 (60.0) | 4 (40.0) | 3.961 (1.118-14.033) | 0.031 |
|  | Colo-rectum/Pancreas/Ovary | 3 | 25 (44.6) | 31 (55.4) | 2.130 (1.258-3.604) | 0.006 |
|  | Lung/Breast/Brain | 3 | 19 (51.4) | 18 (48.6) | 0.612 (0.372-1.007) | 0.057 |
|  | Breast/Ovary/Thyroid | 3 | 11 (57.9) | 8 (42.1) | 2.788 (1.464-5.308) | 0.003 |
|  | Colo-rectum/Breast/Ovary | 3 | 11 (52.4) | 10 (47.6) | 3.631 (1.461-9.024) | 0.007 |
|  | Breast/Uterus/Ovary | 3 | 10 (50.0) | 10 (50.0) | 2.905 (1.234-6.837) | 0.024 |
|  | Head and Neck/Breast/Thyroid | 3 | 10 (66.7) | 5 (33.3) | 2.641 (1.100-6.342) | 0.041 |
|  | Colo-rectum/Ovary/Thyroid | 3 | 10 (50.0) | 10 (50.0) | 5.282 (1.806-15.447) | 0.002 |
|  | Breast/Cervix/Thyroid | 3 | 8 (66.7) | 4 (33.3) | 2.641 (1.100-6.342) | 0.041 |
|  | Pancreas/Cervix/Ovary | 3 | 8 (57.1) | 6 (42.9) | 5.282 (1.591-17.534) | 0.005 |
|  | Cervix/Ovary/Thyroid | 3 | 6 (66.7) | 3 (33.3) | 3.521 (1.222-10.144) | 0.03 |

RR; relative risk, CI; confidence interval
<sup>†</sup> The cancer types regardless of the occurred order
<sup>‡</sup> The age at first cancer diagnosis
\*Relative risk was calculated by comparison of the proportion for those aged <50 years (n=60,648 patients) with two or more cancers, by sex.
There were 3,355 (9.4%) out of 35,508 and 6,905 (27.5%) out of 25,140 male and female patients, respectively, who were diagnosed with cancer
before the age of 50 years. Only statistically significant combinations are reported.

**Figure 2.**
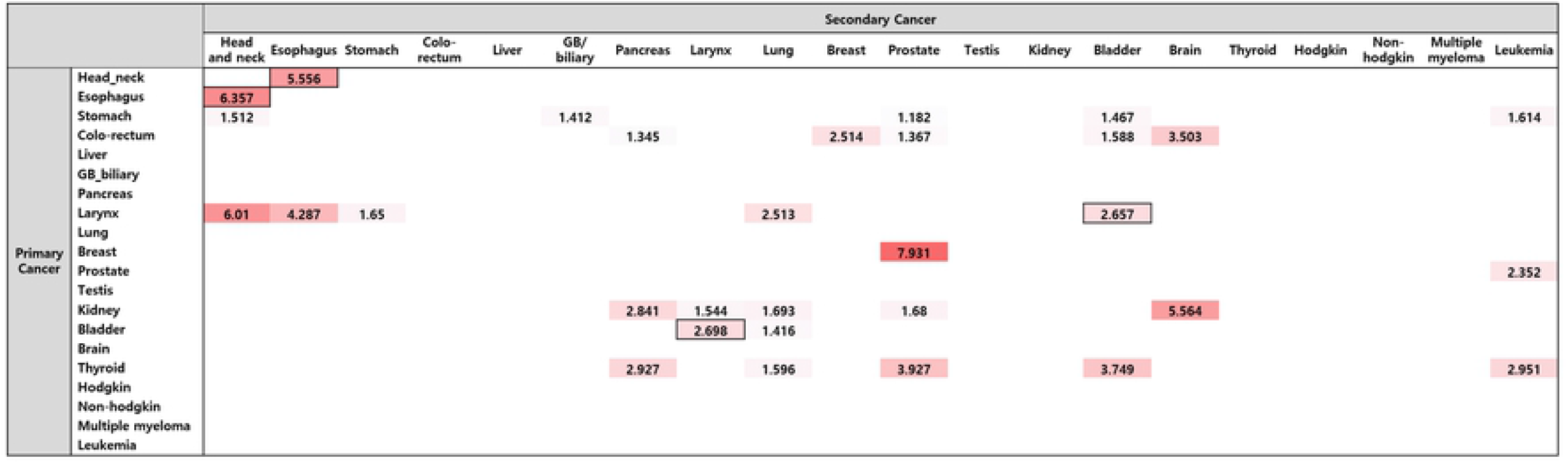
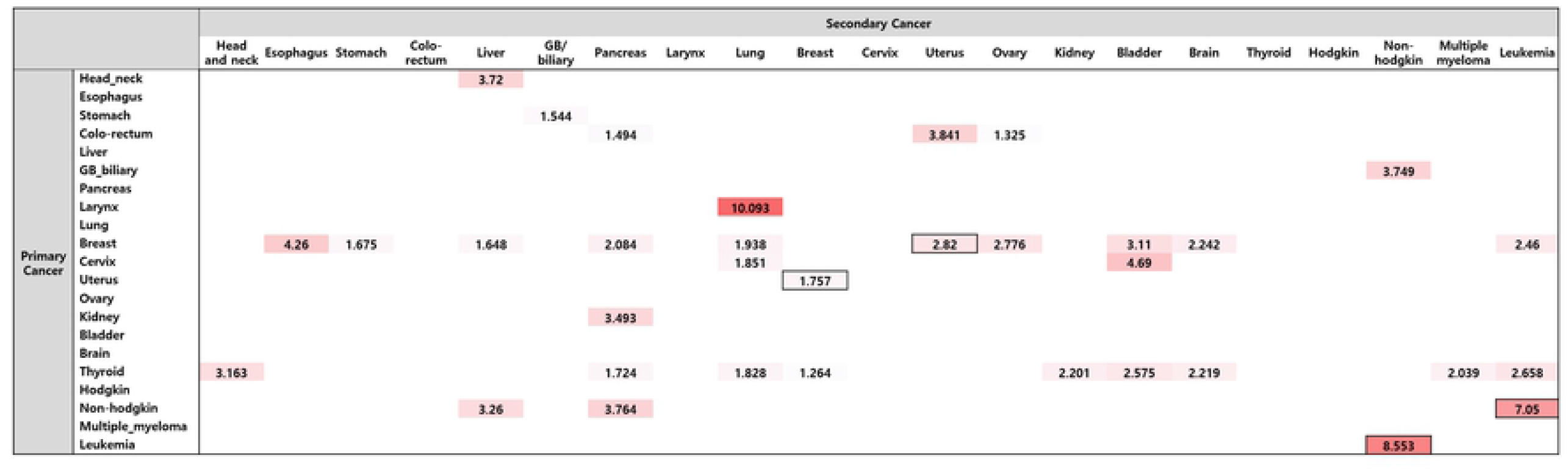
The heatmap for risk of secondary cancers in cancer survivors of A) male and B) female. The number represents age-standardized incidence ratio (SIR) compared to the primary cancer in general Korean population. The intensity of the color represents the SIR and bold square represents the cancer types that were significantly associated with each other regardless of the order of diagnosis. Only statistically significant combinations are presented.

### Susceptibility to HMCS

Table 3 shows the risk of the combination of double cancers in younger individuals, defined as being 10-years younger than the median age at the first cancer diagnosis, by sex. Expectedly, the risk of well-known Lynch syndrome-related double cancers in male patients (stomach and colo-rectum, RR: 1.48, 95% CI: 1.197-1.839, p <0.001) and in female patients (hereditary breast and ovarian cancer syndrome [HBOC]-related cancers [breast-ovary and ovary-breast, RR: 1.70 and 2.38, p = 0.03; p = 0.01, by order of cancers, respectively]) was higher among younger compared to older patients. This suggests that this epidemiologic approach to identify hereditary cancer syndrome, which targeted younger patients with multiple cancers, is reasonable. Some cancer combinations were significantly frequent in younger patients with double primary cancers regardless of the order of diagnosis. The risks of liver-brain, brain-liver cancers, and brain-thyroid, thyroid-brain cancers were higher in male patients (Table 3), while the risk of combinations of breast and cervix, breast and thyroid, cervix and thyroid, and uterus and thyroid were higher in female patients. These combinations could be possible candidates of HMCS; however, the age of onset for secondary cancers might affect this analysis (Table 3 and S1 Table1).

Table 4 shows the risk of cancer combinations in patients with three or more cancers in the younger (<50-years) compared to older (≥50-years) age groups. In male patients, colo-rectum, pancreatic, and stomach cancer, which are Lynch syndrome-related cancers, were mainly observed. There was a higher risk of triple cancers including Hodgkin/non-Hodgkin lymphoma and leukemia in younger patients; however, age could be a confounder in this result because of the younger age at onset of those cancers (S1 Table 1). Triple cancers involving the liver, lung, and brain were frequently observed before the age of 50 years and the combination could be a candidate for novel HMCS because liver-brain and lung-brain cancers also showed high risk in younger patients with double cancers (Table 3). Lynch (colo-rectum, uterus, and pancreas) and HBOC syndrome (breast, ovary, and pancreas)-related cancers were mainly observed in female patients. In addition, thyroid cancer with cancers of the female organs (breast, ovary, and cervix) and colo-rectum were frequently observed, and these could be related to Cowden syndrome caused by *PTEN* germline mutation. Triple cancers involving the lung, breast, and brain were 2.792 times more frequent before the age of 50 years. As combinations of these cancers also showed high risk in younger patients with double cancers, they could be candidates for novel HMCS (Tables 3 and 4). Considering the above results, multiple cancers of the stomach, colo-rectum, pancreas, prostate, liver, lung, and brain in male patients; and colo-rectum, uterus, cervix, ovary, breast, lung, brain, and thyroid in female patients, could be candidates for HMCS.

## DISCUSSION

The present results showed that the risk of secondary cancer was higher among cancer survivors than in those with primary cancers in the general population; thus, careful cancer screening and preventative strategy is necessary for cancer survivors. In addition, common cancers were mainly observed even in cancer survivors, implying that the focus of cancer screening should also be on common cancer types among cancer survivors. However, high-risk secondary cancer types varied by sex and primary cancer type, suggesting that further consideration to tailor cancer screening for cancer survivors is required. For example, lung and bladder cancers were frequently observed as high-risk secondary cancers in cancer survivors of both sexes; and prostate and pancreatic cancer risks were mainly high in male and female survivors, respectively. Therefore, ultrasonography, tumor markers such as prostate-specific antigen (PSA) and cancer antigen (CA)19-9 for prostate/pancreatic/bladder cancer screening, and chest X-ray or low-dose computed tomography for lung cancer screening[21] need to be considered for cancer survivors with specific primary cancers. Based on the present results, a cancer type- and sex-specific national cancer screening program for cancer survivors could be developed and the benefit of new tailored screening programs for cancer survivors needs to be evaluated by further studies.

Strong associations between head and neck/esophagus and larynx/bladder cancers were observed – these are well-known smoking-related cancers.[19, 22] Smoking is one of the strongest carcinogens causing various cancers, and smoking cessation reduces the risk of cancer and improves the survival of cancer patients even after the cancer diagnosis.[23, 24] Therefore, quitting smoking is especially crucial for cancer survivors not only to improve survival but also to prevent high-risk secondary cancers. However, it seems that smoking cessation interventions for cancer survivors have not been successful.[25, 26] Clinicians need to communicate with and educate cancer survivors regarding the high risk of secondary cancer and guide smoking cessation. Considering that smoking cessation services are mainly provided by primary physicians, collaboration between oncologists and primary physicians is essential to provide high-quality comprehensive smoking cessation intervention.[27]

Hereditary cancer syndrome, caused by inherited DNA pathogenic variant in cancer predisposition gene, is related to high-risk and early-onset cancer occurrence, and a half of first-degree family members have a similar risk of cancer. In addition, hereditary cancer syndrome sometimes presents as multiple cancers in individuals. Therefore, the identification of hereditary cancer syndrome is clinically important for both cancer prevention and treatment.[28] The HBOC and Lynch syndrome, harboring germline mutation in *BRCA1/2* and mismatch repair-related genes, are well-known hereditary cancer syndrome related to breast-ovary and various cancers including colon cancer, respectively. The present study showed that multiple cancers in HBOC and Lynch-related sites, breast/ovary/pancreas, and stomach/colon/uterus/prostate/pancreas are frequently observed in younger compared to older groups among patients with multiple cancers. A previous study showed that genetic predisposition is mainly observed in younger rather than older patients with Lynch-related multiple cancers,[10] and HMCS may be mainly observed in younger patients. Therefore, our epidemiologic approach could be a guide to discovering novel hereditary cancer syndrome expressed as multiple cancers, HMCS. We found combinations such as lung, brain, and liver cancers, which are not well-known HBOC and Lynch-related cancers, in young patients with multiple cancers. Because the spectrum of Lynch syndrome is wide,[29] these multiple cancers also could be related to Lynch syndrome; conversely, they could be a novel HMCS caused by other cancer predisposition genes. A whole-genome-wide study targeting patients with these combinations of multiple cancers focusing on younger-age onset is worth investigating.

There is evidence that cancer could be prevented even in hereditary cancer syndrome. Hormonal therapy such as tamoxifen could reduce the risk of breast cancer in patients with *BRCA1/2* mutation carriers.[30, 31] The preventive effect of aspirin to reduce the risk of colorectal cancer in Lynch syndrome has been reported,[32] however, its benefit is unclear for other cancer types. Considering that a recent large genomic cohort study showed that the spectrum of Lynch syndrome is wider than expected,[29] further studies to evaluate the benefit of aspirin for other cancer types should be conducted. Recently, it was reported that a frameshift peptide neoantigen-based vaccine to prevent cancer in Lynch syndrome is tolerated in humans.[33] Therefore, it could be used as a preventive treatment for Lynch syndrome along with aspirin in the future. If hereditary cancer is preventable, early diagnosis of hereditary cancer syndrome may become increasingly important. Developing DNA sequencing technology can make early detection possible at the clinic and popularize the concept of genetic disease.[34] Furthermore, despite the possible disadvantage of genetic screening including false-positives or findings of unknown significance that could increase anxiety and unnecessary clinical examinations, active genetic screening for high-risk patients is still worthwhile.[29] Based on the present study results, patients with multiple cancers, especially younger patients, are good candidates for genetic evaluation, especially candidates with combinations of HMCS.

An interesting association was observed in male breast cancer survivors; they had high-risk prostate cancer as the secondary cancer. The positive association of the two cancers, which has been reported in male patients,[35, 36] could be explained by two hypotheses.[37] 1) It was reported that 4%-40% of male breast cancer patients harbor *BRCA2* germline mutation.[38, 39] *BRCA1/2* mutation causes both breast and prostate cancers; therefore, high-risk prostate cancer in male breast cancer survivors could be related to the germline susceptibility.[40, 41] 2) Most male breast cancers are estrogen receptor-positive, implying that they are good candidates for hormonal treatment; therefore, they can be treated with aromatase inhibitors at the clinic.[36, 42] The suppressed estrogen could cause an imbalance of estrogen and testosterone and hypothetically increase the risk of prostate cancer.[43] This association suggests the necessity of germline testing and careful prostate cancer screening in male breast cancer survivors.

This is the first comprehensive epidemiologic study that covered various cancer types focusing on secondary cancer in cancer survivors. Despite the novel findings, some limitations need to be addressed. The risk of secondary cancer could be under-estimated in cancer survivors who develop cancers with high risk of mortality, such as pancreatic and lung cancers. We could not consider the effect of chemotherapy or radiation therapy that could be related to the incidence of some specific cancers.[44] This is largely attributed to the limitation of accessible data, due to the HIRA policy. We also could not consider family cancer history because the HIRA do not cover the information. Detailed histologic or molecular types of each cancer could not be considered. Some minor malignancies that could have related genetic susceptibility, such as pheochromocytoma and paraganglioma syndrome,[45] could not be evaluated, because they were classified as other cancers (other C00-96) and excluded from the analysis.

## Conclusion

Cancer survivors harbor more high-risk secondary cancers than the general population. Because the high-risk secondary cancers varied by primary cancer type, tailored cancer screening is necessary for cancer survivors. Genetic screening needs to be considered for younger patients with multiple cancers to validate the genetic predisposition.

## Data Availability

All relevant data are within the manuscript and its Supporting Information files.

## Acknowledgements

None

